# The “Trauma Pitch”: How stigma emerges for Iraq and Afghanistan veterans seeking disability compensation

**DOI:** 10.1101/2022.04.11.22273710

**Authors:** Katinka Hooyer

**Author notes:** (KH).

## Abstract

Posttraumatic Stress Disorder continues to be a highly stigmatized disease for the veteran population and stigma continues to be identified as the main deterrent in treatment seeking. Little attention has been paid to how the process of obtaining service-connected disability status can amplify veterans’ perceptions of being stigmatized. The following ethnographic study identified how combat veterans experienced stigma in processing through Veterans Affairs care and the effects of linking a Posttraumatic Stress Disorder diagnosis with disability compensation to perceived stigmas. Stigma was identified in two inter-related areas: 1) the structural level in the Veterans Affairs disability claims process and 2) the individual level in interactions with Veterans Affairs service providers. Results based on veterans’ narratives suggest that the disability claims process, requiring multiple repetitions of personal trauma, coupled with perceptions of institutional stigmas of malingering, created *bureaugenic effects*: a worsening of symptoms caused by bureaucratic protocols intended to help veterans. This process influenced first time treatment users of the Veterans Affairs by deterring treatment-seeking behavior but was not found to affect veterans who had already initiated treatment. Despite the experience of stigma and commodification of their suffering through disability and diagnostic screening, veterans still sought disability compensation. Veterans viewed this compensation as acknowledgment of their loss and validation of their sacrifice.

## Introduction

The convergent relationship between Posttraumatic Stress Disorder (PTSD), veterans’ experiences with PTSD-related stigma, and the need to be diagnosed with PTSD to receive Veteran Benefit Administration (VBA) disability benefits remains under explored. The clear intent of the VBA is to ensure that veterans injured through combat or other military related events receive appropriate financial support, healthcare, and ancillary services to reduce the impact of these events. However, little attention has been paid to how the process of obtaining service-connected disability status can, in fact, intensify the veteran’s perceptions of being stigmatized. This ethnographic study identified how combat veterans experienced stigma in processing through these bureaucratic forms of care and the effects of linking a PTSD diagnosis with disability compensation to perceived stigmas. To fully understand the overlapping nature of these issues for psychologically injured veterans, the process of seeking Veterans Affairs (VA) disability benefits, PTSD, and stigma are explored both separately and in terms of their intersections. While treatment seeking for PTSD among veterans remains a serious public health issue, PTSD also remains the most prevalent compensable mental health disorder in the VBA disability system. PTSD claims have more than tripled in the last decade. About 22% (1,118,041) of all veterans who receive disability compensation through the VBA (4,944,275) have these benefits because they suffer from PTSD [1]. In order to receive disability compensation, a veteran must first go through a series of appointments in the Veteran Health Administration (VHA; i.e. the “VA Hospital”) to be diagnosed and provide evidence that their diagnosis or injury is specifically related to their military service. In the language of the VBA, veterans need to prove that their injury is “service connected”. Some injuries are “conceded”, and military decorations such as the Combat Infantry Badge and the Purple Heart can be considered as evidence of exposure to combat-related stressors. However, these policy changes did not occur until 2010, seven years after the Global War on Terror began. Because of the ethnographic approach taken, this study frames the analysis from two main contexts, the systemic and the individual level. The perspectives of 1) veterans at their local VHA and VBA facilities going to medical and mental health appointments and applying for benefits, and 2) claims officers and healthcare providers working in VHA and VBA bureaucracies, are interwoven on systemic and individual levels. The paper is organized as follows: first, a brief background covering the barriers to PTSD care, stigma and the VA disability claims process is offered along with a consideration of how these three distinct issues interact in psychologically injured veterans; second, the study method is summarized; third, an analysis of findings illustrating how stigma emerges for Iraq and Afghanistan veterans on a systemic and individual level is explored in some detail.

### PTSD and barriers to care seeking in veterans

After over a decade of military conflicts in Iraq and Afghanistan, the number of war veterans diagnosed with PTSD continues to rise. A RAND study conducted in 2008 estimated PTSD prevalence for Operation Enduring Freedom in Afghanistan (OEF) and Operation Iraqi Freedom (OIF) veterans at 13.8% [2]. A recent meta-analysis of PTSD in OEF/OIF veterans found a much higher number, estimating prevalence at 23% [3]. Only slightly more than half of veterans with a PTSD diagnosis seek treatment [4]. According to a Department of Veterans Affairs study, barriers to care include mistrust of the medical establishment, difficulty finding a therapist, uncertainty that treatments will be successful and stigma [5]. Other barriers include career worry [6], high treatment costs [7], negative attitudes surrounding mental healthcare (psychotherapy in particular) and for veterans who are reservists and in the National Guard, the belief that military units will not provide support for mental healthcare [8].

To complicate the situation further, prevalence statistics are based on OEF/OIF veterans who use VA healthcare, yet many veterans do not seek care through this system, instead using civilian providers and insurance [3]. Specific barriers to care for those who do not seek VA services include veteran beliefs that they are ineligible for VA care, prior negative experiences seeking care at the VA, and logistical issues such as distance from a VA facility and available hours for appointments [9].

### Stigma

Stigma, as theorized by Erving Goffman [10], is a process of stereotyping where negative labels (e.g., dangerous, crazy) are attached to a category (e.g., PTSD, veteran), thereby differentiating people as unusual or unacceptable. This “spoiling” of identity results in discrimination, loss of status, and social exclusion. Stigmatization cannot occur without the social power necessary to transform stereotyping into negative consequences [11]. The manner in which the media stereotypes veterans, as threatening and unpredictable, and the ways these representations allow the civilian world to classify the military experience leaves many veterans disconnected to the communities they live in and hesitant to disclose their veteran status, let alone the psychological effects of war [12]. In a health-related context, stigmatized people often hide their condition and forgo treatment. This is especially true when conditions are culturally perceived to be caused by moral transgressions, dangerous, and/or affect one’s appearance (e.g., AIDS/HIV, substance misuse, schizophrenia). Stigma can be so powerful that desired and available treatment is delayed, terminated, or avoided, exacerbating symptoms, and transforming treatable conditions into desperate cases that can result in premature death [13]. Stigma leads to discrimination in housing, employment, social networks, and healthcare [14]. These setbacks create a sense of social defeatism and damage self-worth, cascading into a cycle of increasing stigmatization. Several studies with veterans suggest that general barriers to mental health treatment involve an individual’s fear of being stigmatized as psychologically weak [15-17]. To avoid stigmatization and status loss, many active duty service members do not disclose mental health problems for fear of being perceived as incapable by their officers (chain of command), declared unfit for active service, viewed as unreliable by peers or harming their career [18]. For OEF/OIF veterans with psychiatric diagnoses, stigma and barriers to care were most strongly associated with feelings of embarrassment, perceptions of being viewed as weak, lack of knowledge regarding where to go for treatment and scheduling difficulties [8].

### The disability “claims” process

A military service-connected disability is defined as a disability that is due to injury or illness sustained in or worsened by a veteran’s military service. Within the VA system, it is known as a “service connection” and veterans refer to applying for a service connection as submitting a claim or “claims”. There are eight steps involved in the claims process: 1) claim received; 2) claim under review; 3) gathering of evidence from veteran or medical professional (combined VBA/VHA); 4) review of evidence; 5) preparation for decision; 6) pending decisions for approval; 7) preparation for notification, and; 8) completion and delivery of decision packet [19]. A claims officer assists the veteran in applying for a service connection and claims reviewers gather the evidence needed to access the claim.

The OEF/OIF veterans who are within five years of separating from the military have access to VHA medical benefits. It is a tiered insurance system where veterans who have 30% or more service connection receive free healthcare. This percentage must be maintained to continue to receive these benefits and percentages are re-evaluated periodically. Not all veterans who go to the VHA for care claim a disability connection for reasons discussed later in the paper. Veterans also receive monthly monetary compensation based on their disability rating.

The institutional definition of a disability rating is based on “occupational and social impairment” according to the VA General Rating Formula for Mental Disorders [20]. The VBA bases a rating off a veteran’s ability to work and function in their social world and ratings are made at 0, 10, 30, 50, 70, 90 and 100%. A 0% rating acknowledges there is an illness or injury connected to military service but does not warrant compensation at that point in time. Essentially, a rating is not related to war experience or military trauma but to the *effects* of these experiences.

### The intersection of veteran disability, PTSD diagnosis and stigma

Veterans seek disability compensation for PTSD for a variety of reasons, including material benefit, but also for symbolic reasons linked to acknowledgment for the sacrifices they made during their military service [21]. Disability claims may help the veteran clarify their health issue, offer a recognition of service, and often are undertaken with encouragement of trusted friends or professionals. Veterans deterred from applying for a service connection were concerned with negative public perceptions associated with disability and receiving government aid [22]. Veterans who delay seeking VA treatment are also concerned about the stigma of being labeled mentally ill and the perception of putative responsibility for own their illness given that military service was voluntary [23].

In another study, social perceptions of the undeserving “welfare queen” were found to have negative impact on veterans seeking care and disability benefits [24]. Murdoch et al. [25], found that veterans who received compensation for PTSD had clinically meaningful reductions in symptoms and less poverty and homelessness than veterans who were denied benefits over the course of 10 years. Recent ethnographic studies in psychology [26], sociology [27], and anthropology [8, 28] illustrate that while a PTSD label might be necessary to receive disability benefits for the psychological injuries that military service incurs, the very act of claiming a PTSD diagnosis is often viewed by veterans as claiming individual victimhood, creating a form of moral tension. This conflicts with the core values that military culture embraces, i.e., physical and psychological strength and group loyalty. Military service period may also play a role, Harris et al. [29] found that OEF/OIF and Operation New Dawn (the 2011 Iraq War) veterans who received disability benefits had trouble overcoming self-stigma and alienation compared to other veterans.

These studies typically address stigma as a barrier to treatment and tend to focus on veterans’ individual beliefs, rather than external or structural factors, such as the VBA disability claims process itself. This ethnography helps fill this gap in knowledge by identifying how stigma unfolds for veterans through the disability claims process. The primary objectives were to: 1) identify if and where stigma became a factor for veterans in claiming disability for PTSD, and; 2) the effects of linking a diagnosis to disability ratings and compensation.

## Methods

### Context

The study took place in a Midwestern city at the local VHA medical center, Veterans Benefits Administration (VBA) offices, and three local veterans service organizations sites.

### Ethnographic approach

As a first phase in public health services research, qualitative findings can generate hypotheses that inform larger scale studies. Due to the lack of research in this area, stigma processes could not have been postulated *a priori* for hypothesis testing and an ethnographic approach allowed these processes to emerge throughout the data collection and analysis. This approach specifically seeks to understand the relationship between the micro-level experiences of individuals and macro-level cultural and social systems. Observing, documenting, and analyzing the relationships between these elements are conceptualized as ‘thick description’ [30], especially useful in problem assessment and intervention design in health promotion research and practice [31]. Ethnography further allowed for the identification of the personal meanings of disability ratings for OEF/OIF veterans [32].

This ethnography combined direct, *in situ* participant observations that veterans consented to (accompanying veterans to VHA and VBA appointments), in-depth semi-structured interviews (veterans, disability claims reviewers and VHA mental health providers) and document analysis (VHA screening tools and VBA procedures). Document analysis [33] of participants’ “VA paperwork” including disability determination letters, applications, and disability rating schedules. Veterans shared these documents of their own accord within the context of VA appointments and interviews. Collecting data at these three levels of participant observation, interviews, and documents established confidence in the findings through constant comparison (triangulation) and as a part of theory generation [34]. The ethnography was designed and carried out by a trained qualitative researcher with seven years of experience and in fulfillment of a doctoral degree.

### Participants

Iraq and Afghanistan veterans (n=15; 10 men and five women) participated for 18 months while seeking a VBA service connection for PTSD or having an approved claim. Three additional participant groups were interviewed during this time to gain a broader understanding of institutional practices and perspectives: four claims officers, three VA patient advocates, and seven VA mental healthcare clinicians (e.g., psychologists, psychiatrists, clinical psychiatric nurses). IRB approval was granted by the University of Wisconsin-Milwaukee (IRB# 12.384).

### Recruitment, sampling & participant characteristics

#### Veterans

Veteran recruitment was accomplished through flyers at local veteran service organizations and through snowball sampling [35]. Once the study had 10 participants a purposeful sampling procedure was utilized to obtain more variability on categories of interest and to provide better representation of the OEF/OIF population. Categories of interest included PTSD diagnosis and gender; underrepresented minority groups; military specialty (combat enlisted, combat officer, support enlisted, support officer) and veterans’ military status (separated from military, enlisted in National Guard or Army Reserves). An attempt to mirror the overall demographics of the military in the sample was made. Recruitment continued until 15 participants were enrolled to account for dropout. In qualitative research, to reliably ensure consensus and adequate data collection at least ten interview participants are recommended [36]. Serendipitously, there was great variability in participants’ claims status including: PTSD service connection denied, claims being re-opened, claims granted, new claims pending, and old claims being re-evaluated for an increase in rating.

Four veterans did not complete the study by not returning scheduling calls and one person died from drug withdrawal complications. Of the remaining 10 participants, six had a PTSD diagnosis and a disability rating, while four were in various stages of filing a disability claim. Seven were men and all served in the infantry (combat) and three were women. These women, technically “noncombat” still experienced combat through “beyond the wire” (outside the base) convoys, enemy attacks, and firefights. One veteran was a commissioned officer in the Marines and three were noncommissioned officers in the Army or Army Reserves. Three were of ethnic minority status. Two participants were still working in the military, one in the Army Reserves and another in the National Guard.

One woman in the Army Reserves did not pursue a PTSD diagnosis or service connection because of her leadership status as a noncommissioned officer and fear of losing that status. Notably, a PTSD diagnosis renders soldiers undeployable and the Department of Defense can pull the electronic health records of VHA patients.

### VA Claims Officers and Patient Advocates

These participants were recruited through face-to-face meetings with leaders of veteran service organizations and through their network referrals. Four claims officers and three veterans’ advocates participated. Six of the seven were veterans.

#### VA mental health clinicians

VA clinical leaders were supportive of the research. A project presentation was conducted at a staff meeting to recruit mental health clinicians who worked directly with OEF/OIF veterans.

### Interview procedures

A series of semi-structured interviews were conducted, focusing on the PTSD experience, diagnostic procedures, and compensation process to identify variability in participants’ multiple perspectives. With each of the 10 veterans who finished the study, a series of three interviews administered over 18 months at three to five-month intervals was conducted. For the five veterans who did not complete the study, two completed one interview and three completed two interviews. A total of 38 veteran interviews were conducted. Interviews varied in length and focused on veterans’ thoughts surrounding their diagnosis, their choice to pursue a disability claim and experience in this process, and their experiences with treatment. The 18-month study design allowed for capturing veterans’ experiences as the lengthy bureaucratic process ensued. The VA claims officers, patient advocates and VA providers took part in one interview each, for a total of 14 interviews. All interviews were audio-recorded and transcribed.

### In situ observations of the claims process

To augment the veteran interviews and gain deeper insight into the claims process, direct participant observation was conducted (and consented to) at the local VA medical center and service organizations where the author engaged in informal interactions with veterans and providers. The author was able to follow one woman through her claims process from beginning to end, accompanying her to her initial claims appointment to the final disability and compensation evaluation appointment. This veteran also contacted the author at every touchpoint with the Veterans’ Health Administration (VHA) and the Veterans’ Benefits Administration (VBA). In the ethnographic research process, participants sometimes do things we do not expect. Veterans often called or texted the author, providing updates after interactions with VHA and VBA staff. Other times the author received photos of squad reunions or links to news articles participants thought would be of interest. These observations and interactions were documented through field notes, processed on a computer, and analyzed in qualitative software. Approximately 80 pages of field note data were generated in these observations.

### Analysis

The entire project generated over 700 pages of data; however, this study focused specifically on those segments that addressed stigma and disability, the claims process, and the personal meanings of disability determinations and ratings for research participants. The scope of data was narrowed through qualitative software Dedoose version 8.1.

Analysis began with a review of the data to create overarching codes. These codes described broad common themes in the data such as VA, PTSD experience, and social life. Excerpts of data were sub-coded, under these broad categories, that related specifically to the claims process, stigma, and disability. These sub-codes reflected participants’ experiences. In comparing and contrasting these sub-coded categories across data sets patterns and the relationships between them were identified [37]. The patterns that emerged were notated through analytical memos that were linked to the coded data. In using this constant comparative method, and inductive and deductive analysis, themes and variations of themes were discoverable [38].

As interviewing, observations, and coding proceeded, new codes emerged, and established codes went through revisions. The flexibility of this coding process allowed for new theories regarding veterans’ experiences of stigma and disability compensation to develop [39]. Validity was addressed by: 1) triangulation of interview, observation, and document data; 2) checking emerging theories with research participants, and; 3) only reporting categories that were represented by at least half of participants.

### Findings

#### The Trauma Pitch: stigma on the systems level

> *You’re being put on display having to talk about it every time. Basically, you are being soul raped anytime you have to talk about it*.
>
> Male, OIF veteran and claims officer

In this sample, veterans had recounted their trauma four to seven times in the disability claims process depending on what type of VA facility they started with (an outreach center, the VA hospital, or the VBA regional offices). For example, one veteran started at a Vet Center (a satellite of the VA) with a counselor (1) and then was referred to the VA to ‘get into the system’ in order to file a claim and receive future services. Here he was connected with a service officer (2) who helped him file for disability and set up medical appointments. He told his story again to a primary care provider (3) who made an assessment and referred him to a specialist (4) for a traumatic brain injury evaluation (with medical students observing). After the evaluation he was referred to a psychologist (5) for a diagnosis. Then, he saw a psychiatrist (6) for medication. This veteran gave his trauma account six times to six different people, all of whom needed to know the details of his trauma in order to make an assessment. At minimum veterans see a primary care physician, a disability claims officer, a psychiatrist, a psychologist and finally a compensation and pension (C & P) examiner (not necessarily in that order). A number of veterans also have clinical care coordinators or clinical social workers.

It is this process of retelling their story “multiple times to strangers” where veterans experienced a devaluation of their trauma that, in turn, undermined the veterans’ relationship with treatment.

> *You just get sick and tired of telling your own story, it’s like a business pitch and you have to get it down… It’s like a performance. You act out everything except the event. I feel it detaches you from your memories. They lose meaning and you are a year fresh from it. And they’re (providers) talking about it like it’s out of a textbook and I’m still feeling the memories and experiencing the symptoms*.
>
> - Male, OIF veteran
>
> *Why would you want to talk to so many people about something that you are ashamed of to begin with? They don’t know what it is like there so to them it is a diagnosis. To you it’s your life*.
>
> - Male, OEF/OIF veteran and claims officer

I conceptualize this repetitive retelling of intimate and emotionally laden memories as the “trauma pitch.” In these bureaucracies of care (VHA and VBA) the trauma pitch was a necessary process to obtain benefits, whether it was healthcare or disability compensation. Objectifying memories in this way was painful because it alienated veterans from their service and those they served with.

#### Moral anguish

The perceived moral judgment of medical providers and the shame veterans felt about their trauma made them hesitant to disclose the full details of their war experience. This was despite the understanding that one’s disability percentage is “based on how messed up you are” (Male, OEF/OIF claims officer).

> *I don’t want to admit my most personal disabilities. I’m ashamed of what I did and especially ashamed of how it affected me to the point where I need help and then am labeled*.
>
> - Male, OIF veteran
>
> *I’m not going to tell a total fucking stranger the deep-rooted feeling… how I lost guys… how I killed… that’s part of who you are, you don’t just tell these stories… you don’t talk about traumatic stuff because it brings back bad thoughts and feelings. It’s embarrassing to yourself*.
>
> - Male, OIF veteran
>
> *Even if the person is a health professional, they are still human, they are still going to judge you*.
>
> *-* Male, OEF/OIF veteran and claims officer

#### “Unplanned Exposure Therapy”: e*xacerbating symptoms*

The process of having to tell strangers about their trauma created much discomfort for veterans and in their view, made their symptoms worse. After VHA/VBA appointments veterans expressed feelings of becoming “…withdrawn and want(ing) to be away from the world”, of “…feeling bad about myself”, and “retreating to the basement and not talking to my wife or daughter.” One male veteran explained:

> *… the evidence (of combat-related trauma) isn’t there even though you have been going to the doctor there for HOW LONG?! It pisses you off and creates resentment. There is the anticipation of something bad (happening) again*.

As one female veteran put it, “The stress alone triggers my PTSD. I have to live it all over again… My blood pressure goes up, I start to sweat, I can’t focus. I start thinking about that stuff. I can’t sleep…” Another male veteran stated, “It would ruin my whole week, that one bad interaction. You feel vulnerable after that… I’d go back into my hole. That’s why I don’t go there (VA) anymore.” These feelings that veterans experienced are consistent with the Diagnostic and Statistical Manual of Mental Disorders V [DSM-V] PTSD symptom criterion B4/5, C2, D2/6, E1/5/6 [40] One male Iraq veteran summed the claims process up as “unplanned exposure therapy”.

#### Threatening collective *and personal identity*

In addition to veterans’ experience of symptom aggravation, the trauma pitch placed veterans’ pride and identity at stake, making veterans:

> *…clam up in the compensation and pension exam and as a result get a 10% rating when they deserve a 50% rating*.
>
> - Male, OIF veteran and claims officer
>
> *…the stuff you see and do (at war) is not something you want others to see, people getting killed*
>
> *… you have a persona you have to keep. Pride is a big thing*.
>
> - Male, OEF/OIF veteran and claims officer
>
> *When I’m told to tell my story time and time again, I try to downplay it. I won’t say how bad it was… tell all the details because I don’t feel like telling someone about the people that were killed, you know, dying moments. I don’t feel people deserve to know those moments. Those were my moments. I don’t want people to take that away from me, tell me how to think about it, how to feel about it*.
>
> - Female, OEF/OIF veteran

These “dying moments”, intense personal experiences of war, were not something veterans wanted to corrupt in any way. They hold significant meaning and putting them into words diminished them. Veterans cherished and took ownership of these experiences and although they lost loved ones, they did not want to lose the fragments of their memories, or what it meant “to be a veteran”. And yet, these memories felt like they lost value upon every retelling:

> *Having to tell people over and over makes it fake, not real… it desensitizes you. I don’t want that to happen*.
>
> - Female OEF/OIF veteran

Altering the trauma memories in this way was a dehumanizing process that was experienced as insensitive care and having “to remember” in a non-reverent way felt shameful.

> *It’s a PERSONAL THING, it was terrible and it gets diluted when you keep having to tell people about it. And then it gets mangled by docs. They break it down into clinical terms and that takes away from the experience and the memories. You have honor and respect for people you get deployed with, then it gets turned into some med student project where they are matching up the symptoms with the diagram in their textbook*.
>
> - Male, OIF veteran

#### Transforming memories and objectifying sacrifice

Memories particularly became corrupted in a technical manner through this transformation of personal experience into professional expertise (i.e., “checking boxes”). For veterans, this diagnostic process depersonalized traumatic events by turning them into quantifiable symptoms, scores, and statistics: “I think they downgrade (scores) because they need positive evidence in light of the public criticism, they are receiving… so now I’m a positive statistic” (Male veteran applying for an increase in PTSD disability rating and concerned about his score). One such scale, the Combat Exposure Scale includes questions like: “What percentage of the soldiers in your unit were killed (KIA), wounded or missing in action (MIA)?: (1) None; (2) 1-25%; (3) 26-50%; (4) 51-75%; (5) 76% or more” and “How often did you fire rounds at the enemy?” Depersonalizing experiences in this way contributed to the detachment that veterans already felt.

As experiences became “diluted” through every round of assessments, veterans unanimously expressed anger at the depersonalized bureaucratic process that was required for them to receive compensation. Veterans became angry both when they needed to tell everything in detail in order to get a high rating (they assumed that the VA already had all this evidence in a file) and when the VA awarded them a lower rating than they thought they merited.

The claims process was described as:

> *…very impersonal, I feel like they don’t care*.
>
> - Male, OIF veteran

Another male veteran stated that when he went for his traumatic brain injury (TBI) test:

> *…there were four med students in there, their trainer and my doctor. They were asking me questions and I felt like they were cross-examining me to see if my statement was the same (as my claim) and my story held up*.
>
> - Male, OIF veteran

TBI and PTSD have many of the same symptoms so often veterans will go through screening for both if they have had a head injury or were exposed to a concussive blast.

The continuous and multiple evaluations made veterans not only feel like they had to keep their story straight (difficult and stressful because memory is affected by trauma) but more significantly, it made them feel like they had to defend their reactions to war:

> *I have to justify why I have an issue, why I feel the way I do. It makes you feel bad… I think they are looking for inconsistencies in the story… I go in one day and tell them I saw 5 IEDs (improvised explosive devices) and the next time they ask I say it was 3 IEDS… I betray my own convictions entering that building because I don’t think the VA is in the business of helping veterans at the expense of these providers who are there to help…*
>
> - Male, OIF veteran

### Challenging provider ethos

Providers struggled with the claims process as well, in particular, the need to generate diagnosis rather than treat symptoms:

> *The most painful experience of your life gets turned into a pain-scale and there is something really wrong and invalidating about that… they (veterans) know things about themselves we only know intellectually*.
>
> - VA psychologist 1

Some seasoned providers digressed from these screening tools to offer a more sensitive evaluation. The psychologists in particular felt that the parameters of these tools were very limiting:

> *…certainly I can check off boxes, I can record symptoms, but I feel like that, you know, by doing that you lose a lot of the qualitative richness of each person’s experience… you lose some of these other contributing issues that are outside those check boxes like the guilt, the grief, the changes in identity and world view, and that I really, really, really, hate*.
>
> - VA psychologist 2

### Providers as *victims of bureaucracy*

Veterans often expressed empathy for providers, viewing them as victims of the system. The impersonal nature of the VA was chalked up to “…people were overworked and just going through the motions.” Despite this empathy for VHA mental health providers there was a general distrust and fear of incompetency on the part of VBA, compensation, and pension (“comp and pen”) evaluators, and the rating system in general. One particularly disparaging story sums up the anxiety and anger that the compensation process can generate:

> *She (physician doing the evaluation) wouldn’t even look at me in the face and she was reading my file: “knocked out by IEDs, mortars*.*” I had numerous concussions and she said there was not enough evidence. That’s because I was infantry! Your corpsman (medic) would push out with 40 Marines… that (incident) wasn’t documented because I was not on base! We were getting shot at. She did not understand that. Then I almost had a panic attack because she did not know what an IED was. Here she is, totally ignorant of my injuries. The lady who is in charge of my disability has no idea what’s going on!*-
>
> Male, OEF/OIF veteran

Although VA policies have changed to liberalize the evidentiary standard (to establish the link between trauma and service) as of 2010, stories like this circulate through the “Joe Network” and are difficult for the VA to recover from. These compensation evaluation experiences made some veterans question the competence of VHA mental health providers and contributed to veterans delaying or avoiding treatment and deterring reapplication for rating increases or appealing a denied claim.

### Blue Falcons and frauds: stigma on the individual level

> *The institutional perception of VA is that they (veterans) are coming to the treatment setting with ulterior motives*.
>
> - VA psychologist

Stigma also unfolded on an individual level in the claims process. Some veterans felt as if their evaluators automatically labeled them a fraud and experienced outright stigma in their claims process: “My comp and pen nurse made the comment ‘Who is telling you what to say when coming in for your exam?’ She’s not following the ‘benefit of the doubt’ requirement in the 38 CFR” (this is the VA’s Schedule for Rating Disabilities) (OEF/OIF veteran and VA disability rater). However, many understood the logic behind the system and condemned those veterans that were faking their symptoms for compensation.

> *He (comp and pen examiner) said “Expect 10%, we’ll see you later*.*” (He thought I was) more or less just looking for a handout and unfortunately there are a lot of people in the VA system looking for a handout, which screws everything up for the people who actually need it*.
>
> - Male, OEF/OIF veteran

As one male veteran described those who are fraudulently gaming the system:

> *There’s a military name for them, Blue Falcons, they just care about themselves and don’t think about how their actions affect everyone else. They are the ones who sneak a candy bar in (during training) and the rest of us have to do push-ups while he eats it*.
>
> - Male, OEF veteran

### Proving yourself

Built into the claims process is the need to produce evidence of trauma, or a “verifiable stressor.” Not surprisingly, at the time of this study all the veteran participants felt they had to “prove” they experienced trauma to their individual assessors. In other words, there was a feeling of judgment before the process even started. As a result of having to produce this evidence repeatedly, veterans felt that the VA did not believe them, care about them, or want to compensate them.

> *You feel like you are giving a testimony. I told you the story. It’s on record. It’s the same story. Why do I have to do that for level 3 and 4? It’s intruding*.
>
> - Male, OIF veteran
>
> *Their job is basically to deny you and so they don’t want to know you personally*.
>
> - Male, OIF veteran
>
> It is your responsibility to prove why you deserve compensation. If you don’t get those records to them (even if they have them) they won’t compensate you. They don’t help you. They don’t want to pay… You get so frustrated you just give up.
>
> - Female OIF veteran

This bureaucratic necessity of providing evidence of trauma created anxiety in a majority of participants.

> *It was very nerve-racking… because they ask you so many personal questions and its doctors you don’t deal with on a regular basis, it’s not YOUR doctors (the ones) that actually might care about you. It’s their job specifically to see random people every day. Their decision is what the disability will be based on, your percentage, and that’s a huge deal, and that like really scared*
>
> *me*. - Male, OEF/OIF veteran

The percentage is a big deal because it translates directly to healthcare benefits and financial assistance.

This perspective of veterans, that the VA does not want to compensate them, is in stark contrast to the providers in this study: “Everyone I work with gives the vet the benefit of the doubt. We all hate the comp and pen process. We care about the vets and hate to put them through this” (VA psychologist). At this particular VA medical center, mental health providers were required to do a quota of compensation and pension exams per month. This dual role that crosses clinical and administrative services created conflict for many because it “muddies the water” of treatment as well as the therapeutic relationship:

> *Our lane is clinical. Our lane is not benefits. Do not cross out of that lane and begin to make statements, write letters, tell people ‘you need to be 100 percent certified’. Do not do that. If you do that you are not helping the veteran*.
>
> - VA psychiatrist

### Invisible injuries: “I’m a 25-year-old in 50-year-old body”

So, while there was some provider resentment towards veterans who used them as a tool to game the system, it was intertwined with a genuine concern over the long-term effects of disability compensation on the future potential and livelihoods of young veterans. Yet, a number of veterans felt their injuries (physical more than mental) were not being taken seriously because their age and bodies did not reflect their pain.

> *I feel I get a little shafted when I go to the medical center because they look at me like a 25-year-old kid that should be healthy. And I will complain about “Oh my back is screwed up, this and that” and if an 80-year-old says that they are jumping through hoops trying to get braces trying to do this, this, and this, and when they hear someone like me bitch, I think they think “Oh yeah he is just being a whiner or a complainer, he is just trying to get money*.*”*
>
> - Male, OIF veteran

Even though veterans felt more stigmatized when it came to physical complaints, the interplay of bodily function and physical appearance was linked to PTSD symptoms. For most this played out in their symptoms of hypervigilance, anxiety, and depression. For example, if the body was “jacked” (slang for messed up) then hypervigilance was more difficult to manage, thereby creating more anxiety.

> *I went to the VA last week and they kept making comments of how much I work out because I am a bigger guy and I don’t look like I’m overweight or something. Yeah, I try and take care of my body, I was marine infantry and that was pretty implemented into my life. But I am fucked up, I am jacked. I mean this weekend I could not even turn my head. And it bothered me so much because I feel vulnerable. I feel like I am not 100% so anytime I do a crazy movement, and me being a “crazy veteran” and me always thinking in fighting mode, well…*
>
> - Male, OEF/OIF veteran

### Golden Handcuffs: compensation as counter-therapeutic

Many providers felt that, at some level, compensation hindered treatment through the possibility of incentivizing illness. As one VA psychiatrist explained it:

> *Here’s our problem as VA providers… we are training as mental health providers to help someone who is distressed and sick. The problem is we have people coming in with different motivations - they need to have me writing in their chart… to convince them (VBA) that ‘I am sick so I can get my paycheck and you are a tool that is going to help me with that… I need a letter from Dr. L telling me that I am really sick, and I can’t work anymore’. That would be counter-therapeutic for me to do that… I have been used sometimes and it bothers me to this*
>
> - VA psychiatrist, OEF/OIF veteran

Providers wanted what was best for their veterans, but many felt conflicted in encouraging disability compensation, also referred to as the “golden handcuffs.” As one clinical nurse put it:

> *I will be really honest with you, that’s (compensation) a huge struggle for me*… *With all of my heart I want to get people into the system and I want to get them connected with every benefit they deserve, however… (you) have a 22 or 23 year-old sitting in your chair and then you are kind of like giving them all these things (benefits)… up front the need is there, believe me, I have worked with plenty of people who come home broken, have a mortgage payment, they have a family, they can’t get a job due to symptoms and… so of course we get them compensated to the highest level possible that we can to help them in that area, but yet then… where is the incentive to get them into a different mindset? Again, when I have people sitting there in my office and I am working with them and I develop that relationship, I always find myself asking them out loud you know, what were your dreams when you were a kid?*

These 10 interrelated areas identified above describe how stigma is experienced by veterans seeking out a disability determination. The stigma experiences that emerged during the VA claims process, from the veteran perspective, were entangled in bureaucratic processes at a systems level but also at an individual level with providers (Fig 1).

**Fig 1:**
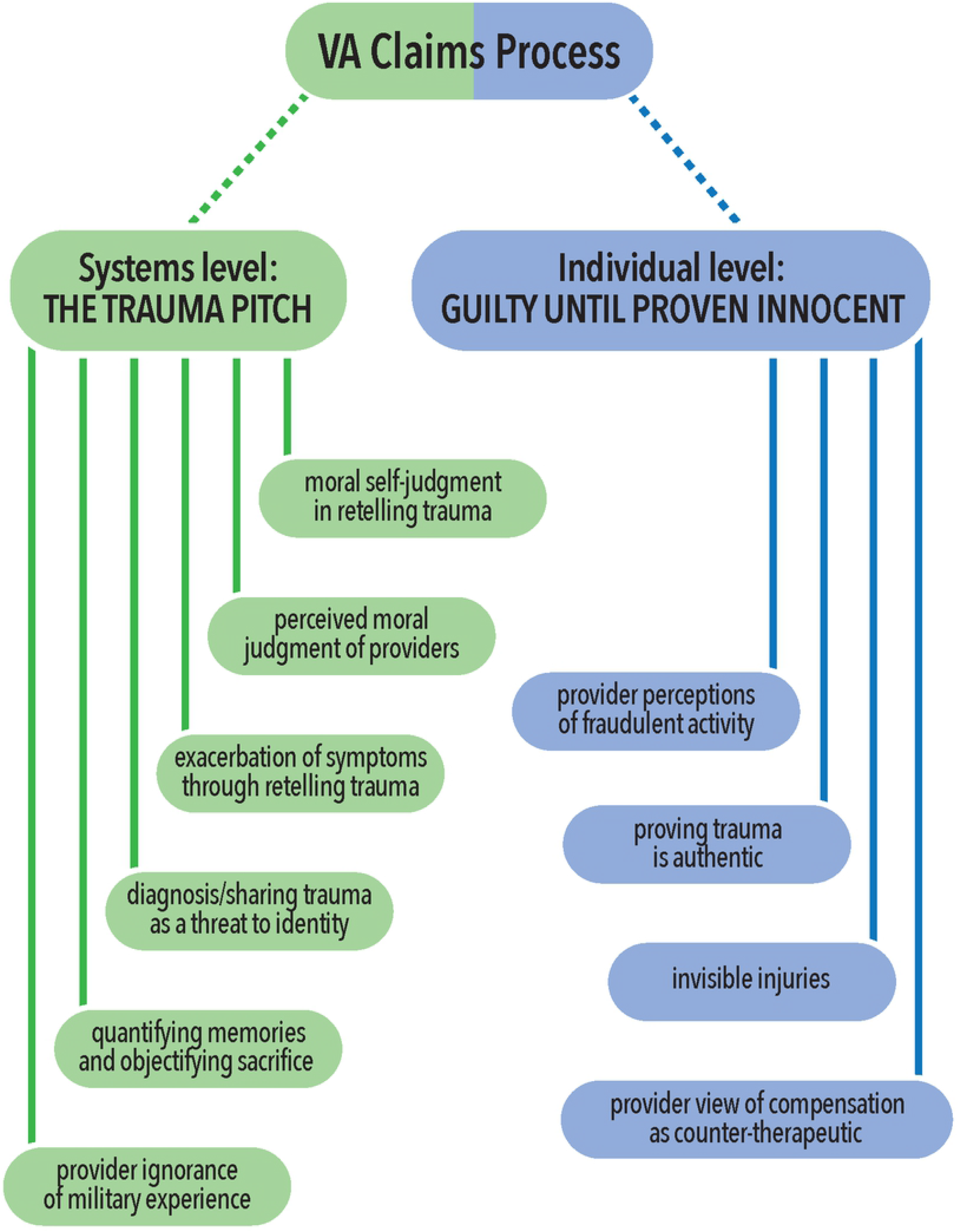
Veterans stigma experiences in seeking PTSD disability determination.

## Analysis and discussion

I have identified 10 interrelated areas that contributed to stigma experiences in the VA disability determination process from the veteran perspective (Fig 1). The trauma pitch is at the core of stigma processes on the systemic level (Fig 1) and conceptualized as the many appointments and screenings veterans go through in seeking a disability rating, requiring them to authenticate and repeatedly tell their intimate trauma experiences to “strangers.” These multiple screenings led to an inadvertent exacerbation of the PTSD symptoms of anxiety, alienation, depression, and anger, and were brought on by bureaucratic processes. These negative and unintended side effects of the disability determination process, designed to help veterans, ended up causing more harm than good for the veterans in this study. I have termed this process, bureaugenic effects. Like iatrogenic effects in medicine, (the adverse outcomes that are the result of a medical treatment) bureaugenic effects are the adverse outcomes of bureaucratic processes intended to help veterans. The bureaugenic effects of the disability claims process increased PTSD symptoms and the experience of stigma in this study (Fig 2). Coupled with an institutional stigma of malingering, veterans felt that clinicians and evaluators did not believe their trauma-related symptoms. The process made veterans feel they were being morally judged. While these experiences, and most certainly the outcomes, were real to veterans, at the heart of this issue was an unawareness of the bureaucratic process on the part of the veterans and an unawareness of the negative impact of these processes on the part of the VHA and VBA.

**Fig 2.**
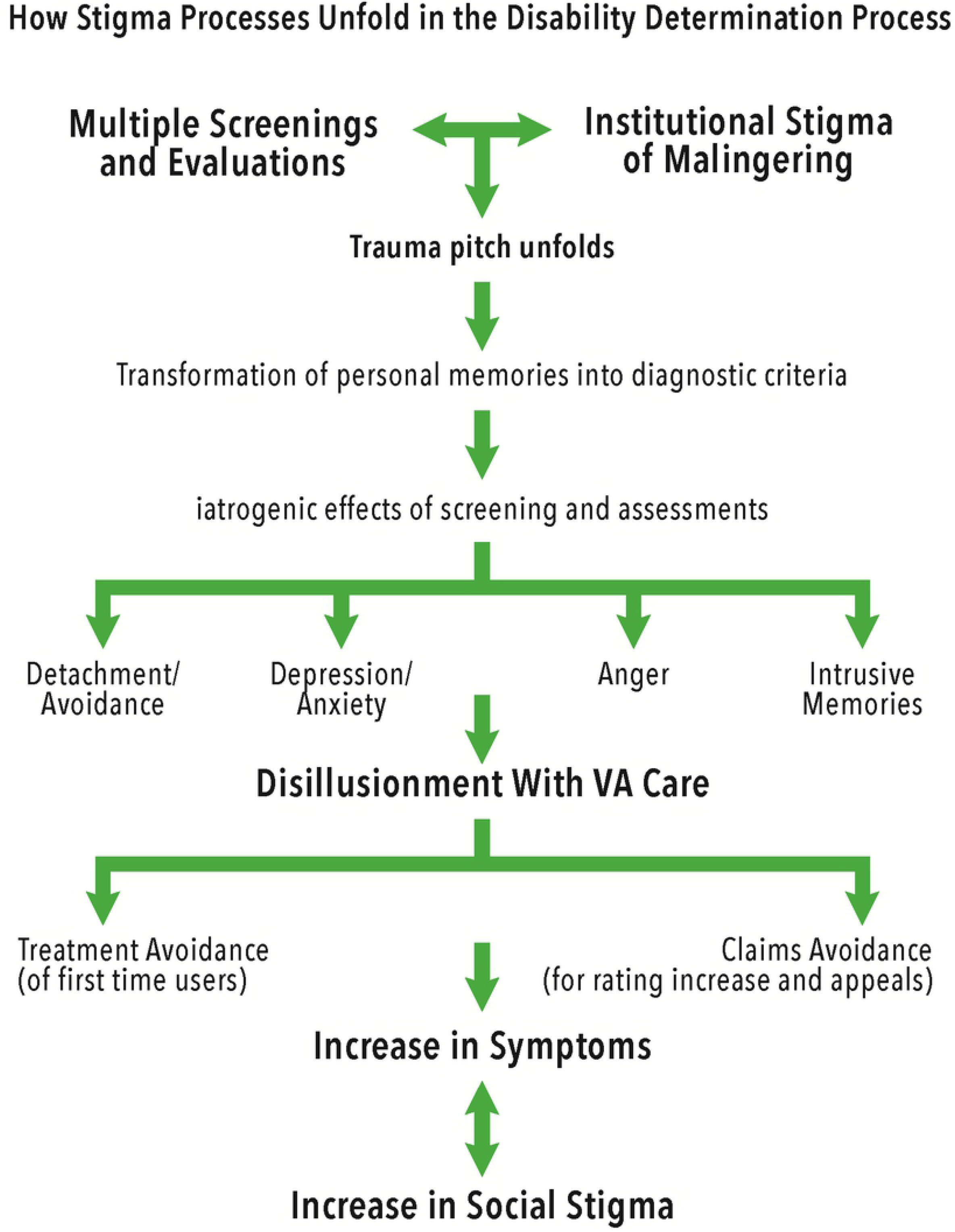
The bureaugenic effects of the disability determination process.

### Bureaucracies of care

The VA bureaucracy is complex. First of all, the VHA is separate from the VBA, a split that most veterans are not aware of. Veterans saw health care providers through the VHA for a diagnosis and comp and pen evaluators through the VBA for their disability rating. To muddle things more, these compensation and pension evaluators were also VHA providers, which in turn created conflict for their therapeutic practice. These two settings, the clinical and the administrative, are co-mingled, and veterans thought all their appointments were part of one process.

Confusion was compounded when veterans thought they were at the hospital for a therapy intake: three of the ten participants who finished the study did not even realize they were going through the disability claims process, having initiated contact with the VHA to start treatment. These veterans were automatically “put into the system” which included application for a disability rating. One of the participants in this study received a “surprise” check of over $3,000 in back pay for a claim he did not even know he started. As one provider put it, “At this point they want a listening ear and instead someone is typing away at their computer.”

At the time of this study, veterans did not know what the status of their claim was for up to ten months. Improvements have been made since this study was completed as veterans can now track their claim electronically to see where it is in the process by registering for eBenefits at www.ebenefits.va.gov. Veterans can also visit VA’s ASPIRE web site where average processing days for specific state regional VBA offices can be assessed.

All claims personnel felt it was critical for veterans to have a service officer lead them through the disability claims process, “They should not attempt this bureaucratic process on their own.” All VHA and VBA staff acknowledged difficulties in navigating the system, especially for those veterans with PTSD who are already contending with anxiety, anger, difficulties focusing and depression. A few veterans related that they had abandoned the process multiple times.

### Meanings of disability ratings

In addition to lack of awareness of the bureaucratic process, there also existed a significant diversity in the meanings of disability ratings between veterans, service providers and the VA. It was this gap in veterans’ personal understanding of ratings and broader institutional definitions where stigma emerged. For example, from the veteran point of view, a disability rating was often a validation of their war trauma. The rating provided a stamp of truth in a system, they believed, was designed to catch frauds. A disability rating was viewed as a form of acknowledgement for their suffering and something they “deserved” for their sacrifice.

In contrast, from a clinician perspective, “claims is not about validation, treatment is” and disability compensation is about “what you can and cannot do” (related to functioning in society). As a result of this difference in meaning, veterans went through the claims process feeling that their trauma had not been recognized, and often felt their ratings were unfair and inaccurate.

Moreover, veterans felt that they were being rated for their experiences (e.g., how many deployments they went on, how many times they were blown up, etc.). This is understandable considering the tools used to screen for PTSD such as the Combat Exposure Scale that quantify and translate war experiences to a percentage. But the institutional definition of a disability rating is not about war experiences, it reflects the effects of these experiences on “occupational and social impairment.” This is what the VBA bases a rating off of according to the VA General Rating Formula for Mental Disorders (known as the 38 CFR Book C).

### Social vs. functional disability

Linked to conflicts of meaning, on a systemic level, an ambiguity between the meanings of impairment and disability and their relationship to a rating emerged. The Post Traumatic Stress Disorder General Rating Formula for Mental Disorders used at the VBA rates for impairment that affects the performance of *occupational tasks*. Certainly, someone can be severely impaired but not disabled: in other words, still able to work. This was evidenced by some veterans who with 50-90% ratings (moderate to highly impaired) were able to hold down full-time jobs. As one provider put it, “The reason for compensation got lost somewhere because the idea is that if you are 30% service connected, 30% of your earning potential has been eaten away.” Earning potential is ambiguous also. Theoretically it might be reduced through no longer being able to live out one’s dream. For example, one veteran planned on becoming a physician, but as a result of war injuries, now works in low-level administration. To push the question of impairment and disability further, veterans could be seriously impaired in their social lives, with certain types of employment still manageable.

### Negative impact of screening tools

This study suggests that the screening tools used for PTSD can have a negative impact on some veterans through quantifying and devaluing their trauma. Clinicians felt the screening tools left out important layers of PTSD experience such as guilt and shame and that the diagnostic criteria were too limiting. The general consensus was that PTSD could present with only a few severe symptoms and in “splitting” experience into symptom criteria and “checking boxes” a formal diagnosis was not established. A PTSD diagnosis was viewed as a “communication tool” but not a clinical tool. In other words, it was a way to communicate with the VBA or other care providers what was going on, but secondary to work therapeutically with specific symptoms and behaviors to improve quality of life. These findings are similar to those of Jackson, et al. [41], which found that 59% of clinicians rarely or never use screening tools with only 17% routinely using them, with less experienced providers using them more frequently. Overall, the providers in the present study expressed much frustration over their role in the disability determination process, which in their professional view, conflicted with their training and therapeutic goals.

### Limitations

This study has a number of limitations. It was conducted in a Midwest state in an urban area with a participant sample that was mostly of European decent. A majority were combat veterans and one third were deployed for their first time at the beginning of the Iraq war. This means, for one third of this sample, their first contact with VA was at a time when VA was not prepared for a young cohort of veterans (with unique needs than VA was accustomed to serving). Also, early deployment generally equates to more trauma exposure, symptom severity, and discomfort interacting with providers and institutions. A number of the veterans in this study had first contact with VA before more lenient policy changes surrounding verifiable stressors was instituted (in 2010). As such, these formative results may not be generalizable to diverse veteran populations.

Self-report biases may have been a limitation due to the nature of the topics explored, as well as the population of study, who tend to be proud in presenting themselves. However, by including three phases of interviews over 12 months, this research design attempted to mitigate these biases. Recollection bias was a limitation in that memory loss itself is a symptom of PTSD.

## Conclusion

All too often, we think of bureaucratic red tape as the daunting task of filling out paperwork, following excessive protocols, or the process of navigating various low-level rules that frustrate and anger us. For vulnerable populations, like veterans, the impacts on mental health can be far more detrimental. In this ethnographic study, I have identified how bureaucratic forms of care created stigma for 10 combat veterans seeking treatment and disability compensation for PTSD. This study elucidated the multi layered process veterans navigated to qualify and receive compensation for combat PTSD. Ten interrelated areas that contributed to veterans’ stigma experiences in the VA disability determination process were identified. For those veterans seeking treatment for the first time, they were deterred by their perceptions of being labeled as a malingerer and having to prove their trauma. The difficulties in navigating this complex system were acknowledged by both VBA and VHA staff, and clinicians felt that their involvement in the disability determination process conflicted with therapeutic goals.

The VA disability claims process, which required multiple recounting of personal trauma and experienced by veterans as a “trauma pitch” not only worsened veterans’ perception of being judged and stigmatized, but inadvertently exacerbated PTSD symptoms of anxiety, alienation, depression, and anger. The increase of these symptoms brought on by institutional processes are conceptualized in this study as *bureaugenic effects*. In addition to veterans’ experience of symptom aggravation, the bureaugenic effects of the trauma pitch placed veterans’ identity at stake: military values of group loyalty were threatened through the objectification of their sacrifices. This developed through the screening tools that quantified traumas and calculated percentages of disability, placing a monetary value on loss. Paradoxically, many viewed this commodification of suffering via disability compensation as a validation of those losses and sacrifices.

This ethnographic study brings to light the need for institutions to be aware of how their bureaucratic processes impact the people they serve, and how stigma and ill-being are propagated through these processes.

## Data Availability

All relevant data are within the manuscript and its Supporting Information files.

## Acknowledgements

Many thanks to Paul Brodwin, PhD, Zeno Franco, PhD, Chris Antczak, and Jeff de Los Santos who were integral to this research and manuscript development.

The opinions and conclusions expressed are solely those of the author and do not represent the opinions or policy of Policy Research, Inc., SSA or any other agency of the Federal Government. Correspondence concerning this article should be addressed to Katinka Hooyer

## Notes

### Competing Interest Statement

The authors have declared no competing interest.

### Funding Statement

This research project was funded by Policy Research, Inc. as part of the U.S. Social Security Administration’s Improving Disability Determination Process Small Grant Program funding in [7/2012-7/2013], specifically awarded to KH. https://www.prainc.com/ardraw-program-blossoms/ The funders had no role in study design, data collection and analysis, decision to publish, or preparation of the manuscript.

### Author Declarations

IRB approval was granted by the University of Wisconsin-Milwaukee (IRB# 12.384).

